# Genome-wide association study of proteomic aging reveals shared genetic architectures with longevity, early life development, and age-related diseases

**DOI:** 10.64898/2025.12.03.25341518

**Authors:** M. Austin Argentieri, Robert Loughnan, Ying Wang, Calwing Liao, Baihan Wang, William Cheng, Robert Ye, Franjo Ivankovic, Najaf Amin, Canqing Yu, Jun Lv, Liming Li, Neil Wright, Chun Chieh Fan, Aarno Palotie, Derrick Bennett, Zhengming Chen, Alexandra Alvergne, Cornelia M. van Duijn, Benjamin M. Neale, Mark J. Daly

**Affiliations:** Analytic and Translational Genetics Unit, Massachusetts General Hospital, Boston, MA, USA; Program in Medical and Population Genetics, Broad Institute of MIT and Harvard, Cambridge, MA, USA; Stanley Center for Psychiatric Research, Broad Institute of MIT and Harvard, Cambridge, MA, USA; Nuffield Department of Population Health, University of Oxford, Oxford, UK; Center for Multimodal Imaging and Genetics, J. Craig Venter Institute, La Jolla, CA, USA; University of California San Diego, La Jolla, CA, USA; Department of Epidemiology and Biostatistics, School of Public Health, Peking University, Beijing, China; Peking University Center for Public Health and Epidemic Preparedness and Response, Beijing, China; Key Laboratory of Epidemiology of Major Diseases (Peking University), Ministry of Education, Beijing, China; Population Neuroscience & Genetics Center, Laureate Institute for Brain Research Tulsa, OK, USA; Institute for Molecular Medicine Finland (FIMM), HiLIFE, University of Helsinki, Helsinki, Finland; ISEM, University of Montpellier, CNRS, IRD, Montpellier, France

## Abstract

There is still relatively little known about the genetic underpinnings of proteomic aging clocks. Here, we describe a genome-wide association study of proteomic aging in the UK Biobank (n=38,865), identifying 27 loci associated with participants’ proteomic age gap (ProtAgeGap). ProtAgeGap exhibits a strong genetic correlation with longevity (rg = −0.83), and in FinnGen a ProtAgeGap polygenic score (PGS) was associated with significantly increased odds of achieving longevity (n=500,348; OR = 1.43). Additional PGS analyses in All of Us (n=117,415), China Kadoorie Biobank (n=100,640), and ABCD Study (n=5,204) demonstrate reproducible associations across biobanks of ProtAgeGap PGS with obesity, cardiometabolic disease, and osteoarthritis in adults, and with developmental timing in children. Finally, colocalization analysis identified *FTO* as an obesity-related mechanism uniting diverse aging traits. Our results demonstrate a shared genetic architecture across the life course of ProtAgeGap with longevity, early developmental biology, and cardiometabolic and musculoskeletal diseases.

## Introduction

Aging is a complex and dynamic biological process that arises from the cumulative effects of genetic and environmental factors acting across the lifespan. While chronological age serves as a convenient metric of time lived, it fails to capture the substantial heterogeneity in biological aging rates across individuals. Measures of biological age (i.e., biological age clocks) can capture the level of biological functioning of an individual in comparison to an expected level of functioning for a given chronological age to more accurately represent an individual’s functional status and risk for age-related diseases.^1^ Various biomarkers, including DNA methylation-based epigenetic clocks, telomere length, and metabolomic signatures, have been proposed as measures of biological age. However, emerging evidence suggests that proteomic profiling may provide a particularly powerful and dynamic readout of systemic aging.^2^ The plasma proteome integrates signals from multiple tissues and organs, reflecting ongoing physiological processes that contribute to aging and disease.

We previously developed a proteomic age clock using data from 45,441 participants in the UK Biobank (UKB).^2^ Beyond its strong age prediction (Pearson r = 0.94), the deviation between proteomic age from this clock and chronological age (termed proteomic age gap, ProtAgeGap) was significantly associated with the incidence of 18 major chronic diseases, multimorbidity, and all-cause mortality, suggesting that deviations between proteomic age and chronological age may capture meaningful biological differences in aging trajectories. Despite the promise of ProtAgeGap as a biomarker, the genetic determinants of proteomic aging remain largely unexplored. Unraveling the genetic architecture of proteomic aging could provide fundamental insights into the biological pathways that shape inter-individual differences in aging rates and disease susceptibility. Furthermore, genetic risk profiling could help identify individuals predisposed to accelerated proteomic aging, and identifying genetic signals associated with proteomic aging can help to strategically leverage biobank data to connect ProtAgeGap with other developmental and longevity phenotypes in large-scale population cohorts where proteomic data have not yet been generated in large numbers of samples.

Here, we present one of the first genome-wide association studies (GWAS) of proteomic aging, leveraging genomic and proteomic data from 38,865 European ancestry participants in the UK Biobank. We constructed polygenic scores (PGS) for proteomic aging and systematically tested their associations with thousands of disease outcomes in the FinnGen (n=500,348), All of Us (n=150,000), and China Kadoorie Biobank (CKB; n=100,640) cohorts, and with developmental and puberty traits among adolescent participants from the ABCD Study (n=5,204). Our results provide further insights into the biological pathways underpinning proteomic aging, and through leveraging genetic analysis we illustrate the shared biology between proteomic aging and a broad landscape of clinical endpoints, aging-related traits, and longevity.

## Results

### Phenotypes

Characteristics of participants across all cohorts are shown in Table 1. For initial GWAS analyses, we considered UKB participants who were randomly selected for plasma proteomic profiling (n=45,441). Although age at time of blood draw showed differing distributions across genetic ancestry groups among this subset of UKB participants, the distribution of ProtAgeGap showed much more stable distributions across genetic ancestry groups (Fig. 1a). Nonetheless, due to small sample sizes in all non-European ancestry groups, the final GWAS sample only included European ancestry participants (n=38,865; 54% female, age range 40–71 years). Genetic ancestry mapping was used from our previous work in the UKB.^3^

**Fig. 1.**
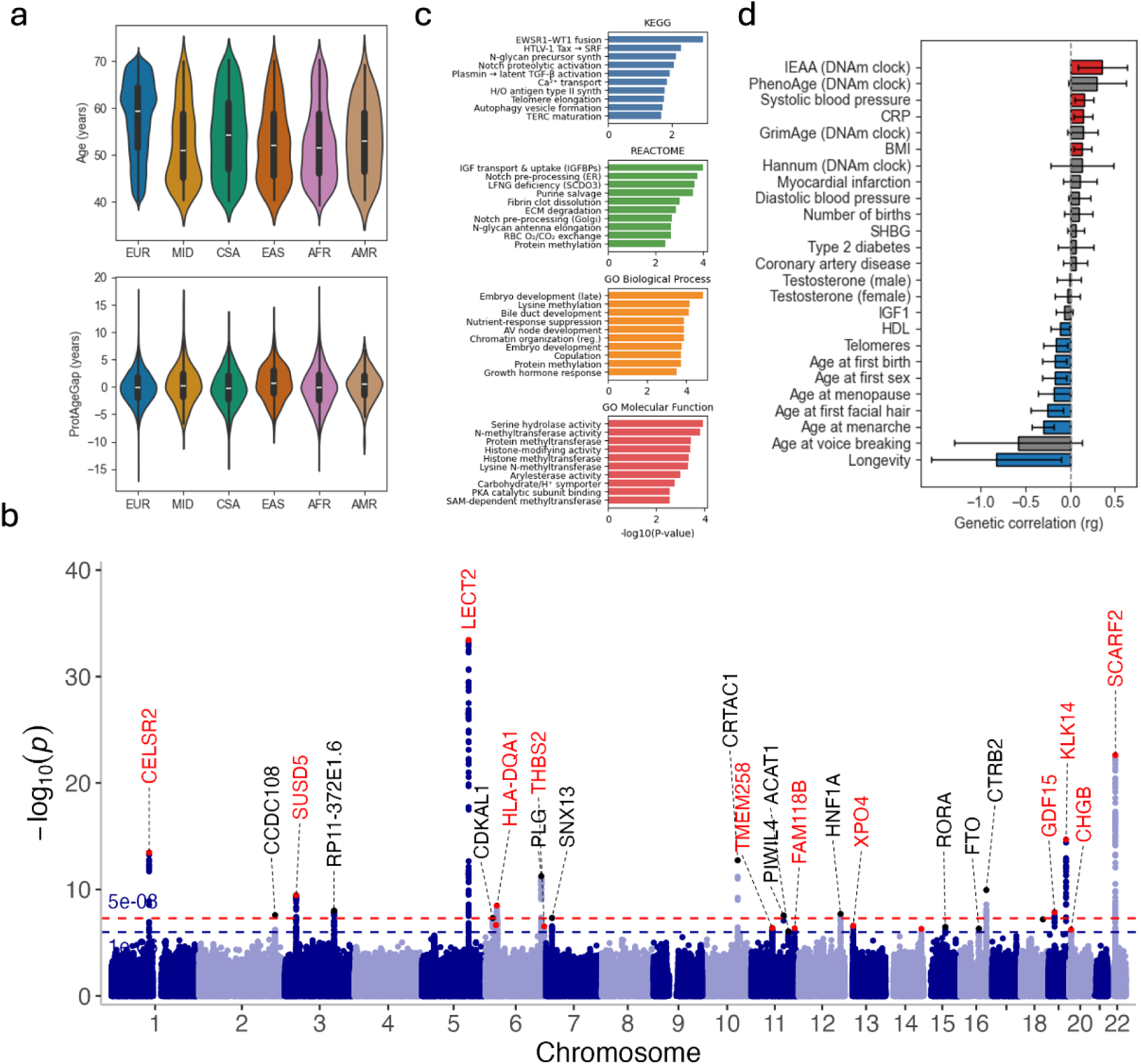
Genetic architecture of proteomic aging. **a)** Distribution of age at blood draw and ProtAgeGap by genetic ancestry in n=45,445 UK Biobank participants. **b)** Manhattan plot from a GWAS of ProtAgeGap in n=38,865 UK Biobank Europeans. Genes shown in red depict variants that are cis-pQTLs for one of the 204 proteins in the ProtAge clock. **c)** Gen set enrichment results from the ProtAgeGap GWAS. **d)** Genetic correlation coefficients (rg) and standard errors for the genetic correlation between ProtAgeGap and aging-related traits. GWAS: genome-wide association study; EUR: European; MID: Middle Eastern; CSA: Central / South Asian; EAS: East Asian; AFR: African; AMR: Admixed American / Latino; ProtAgeGap: proteomic age gap.

**Table 1.** Characteristics of study cohorts.

| Table 1. Characteristics of study cohorts |  |  |  |  |
| --- | --- | --- | --- | --- |
| Cohort | Mean age (SD; range) | Sex | Mean BMI (SD) | Sample size |
| UK Biobank | 57.8 years (8.1; 40-71) | 54% female | 27.4 (4.8) | 38,865 |
| FinnGen | 53.1 years (17.9; 0-104) | 56% female | 27.3 (5.5) | 500,348 |
| All of Us | 59.9 years (16.7, 19-107) | 59.1% | 29.12 (6.7) | 116,272 |
| China Kadoorie Biobank | 53.70 years (11.00; 30-80) | 57% female | 23.66 (3.49) | 100,640 |
| ABCD Study | 12.33 (2.3; 9-18) | 47.2 female | 20.07 (4.68) | 6,598 |

Placeholder for descriptive stats table. In all cohorts, age refers to participant age at the time of blood draw for genetics.

### Genetic architecture of proteomic aging

Using imputed genotype dosages, we assessed common genetic variant (MAF > 0.01) associations with participants’ ProtAgeGap in the UKB, adjusting for age, age^2^, sex, age*sex, age^2^*sex, genotype array, and 20 genetic principal components to account for population structure. SNP-based estimates of heritability using LD Score Regression (LDSC) were h^2^ = 0.10 (SE: 0.02). We found no evidence for genomic inflation (λ = 1.09).

We identified 16 genome-wide significant loci (p<5e-8) and 27 suggestive loci (p<1e-6) associated with proteomic aging, including variants in *CELSR2*, *CTRB2*, *LECT2*, *SCARF2*, *FTO*, and the HLA region that have been previously associated with parental longevity, fat mass, lipids, diabetes, heart disease, inflammation, liver/kidney function, and IGF-1 (Fig. 1b; Table 2). Among these significant loci, we used Sum of Single Effects (SuSiE) to fine-map causal variants, identifying 21 variants with a possible causal influence on ProtAgeGap. We further assessed whether these 21 variants were cis-pQTLs (protein quantitative trait loci) within 1 Mb from the gene encoding for any of the 204 proteins used to calculate the ProtAgeGap score, which would indicate a variant not necessarily driving the overall ProtAgeGap phenotype but rather driving the expression of a single protein within the composite ProtAgeGap phenotype. Only 11 of the 21 fine-mapped ProtAgeGap GWAS variants were a cis-pQTL for any of the 204 ProtAgeGap proteins, indicating that the remaining 10 fine-mapped variants are plausible drivers of the overall ProtAgeGap phenotype.

**Table 2.** ProtAgeGap GWAS significant results.

| Table 2. ProtAgeGap GWAS significant results |  |  |  |  |  |  |  |  |  |
| --- | --- | --- | --- | --- | --- | --- | --- | --- | --- |
| rsID | Chr:Position | Candidate/<br>Closest Gene | Alleles<br>(EF/OA) | MAF | Beta | SE | P-value | ProtAge<br>cis-pQTL | cis-pQTL<br>Gene(s) |
| rs660240 | 1:109817838 | CELSR2 | C/T | 0.78 | 0.17 | 0.02 | 3.30E-14 | TRUE | PROK1,<br>CELSR2 |
| rs56007204 | 10:99625319 | CRTAC1 | T/C | 0.21 | -0.17 | 0.02 | 1.81E-13 | FALSE |  |
| rs11020842 | 11:94317202 | PIWIL4 | A/G | 0.04 | 0.28 | 0.05 | 2.71E-08 | FALSE |  |
| rs102274 | 11:61557826 | TMEM258 | C/T | 0.35 | -0.1 | 0.02 | 4.11E-07 | TRUE | SCGB1A1 |
| rs1613078 | 11:126124835 | FAM118B | A/C | 0.45 | -0.09 | 0.02 | 4.60E-07 | TRUE | CDON,<br>ACRV1 |
| rs4754296 | 11:108006182 | ACAT1 | G/T | 0.14 | 0.13 | 0.03 | 8.94E-07 | FALSE |  |
| rs1183910 | 12:121420807 | HNF1A | A/G | 0.31 | 0.11 | 0.02 | 1.97E-08 | FALSE |  |
| rs11147883 | 13:21444538 | XPO4 | T/A | 0.67 | -0.1 | 0.02 | 2.61E-07 | TRUE | IL17D |
| rs12882815 | 14:101183142 | - | G/C | 0.68 | 0.1 | 0.02 | 5.02E-07 | TRUE | DLK1 |
| 15:60889830_T<br>AC_T | 15:60889830 | RORA | T/TAC | 0.38 | 0.1 | 0.02 | 3.28E-07 | FALSE |  |
| rs72802342 | 16:75234872 | CTRB2 | A/C | 0.08 | 0.23 | 0.04 | 1.12E-10 | FALSE |  |
| rs11642015 | 16:53802494 | FTO | T/C | 0.4 | 0.1 | 0.02 | 4.82E-07 | FALSE |  |
| rs8089764 | 18:64355209 | - | G/T | 1 | -369.75 | 68.36 | 6.37E-08 | FALSE |  |
| rs867192 | 19:51583426 | KLK14 | T/G | 0.2 | -0.19 | 0.02 | 2.06E-15 | TRUE | KLK14,<br>KLK4,<br>KLK8,<br>KLK3,<br>KLK7 |
| rs12710316 | 19:18487541 | GDF15 | T/C | 0.13 | 0.16 | 0.03 | 1.35E-08 | TRUE | GDF15,<br>NCAN,<br>INSL3 |
| rs571026507 | 2:219884769 | CCDC108 | GT/G | 0.11 | -0.18 | 0.03 | 2.52E-08 | FALSE |  |
| rs446659 | 20:5897778 | CHGB | C/G | 0.35 | -0.1 | 0.02 | 5.85E-07 | TRUE | CHGB |
| 22:20776973_A<br>ACC_A | 22:20776973 | SCARF2 | A/AACC | 0.8 | 0.24 | 0.02 | 2.37E-23 | TRUE | SCARF2 |
| rs11706300 | 3:33203298 | SUSD5 | A/G | 0.85 | -0.16 | 0.03 | 3.44E-10 | TRUE | SUSD5 |
| rs2649726 | 3:142694861 | RP11-372E1.6 | T/C | 0.62 | 0.11 | 0.02 | 9.84E-09 | FALSE |  |
| rs31517 | 5:135287029 | LECT2 | C/T | 0.63 | -0.24 | 0.02 | 3.73E-34 | TRUE | CXCL14,<br>LECT2 |
| rs4252165 | 6:161161559 | PLG | A/C | 0.29 | 0.14 | 0.02 | 5.62E-12 | FALSE |  |
| rs115566240 | 6:32601718 | HLA-DQA1 | T/C | 0.24 | -0.13 | 0.02 | 3.27E-09 | TRUE | AGER,<br>TNXB |
| rs9295478 | 6:20716253 | CDKAL1 | A/G | 0.22 | 0.12 | 0.02 | 4.92E-08 | FALSE |  |
| rs28749103 | 6:29903846 | HLA-U | G/A | 0.16 | -0.14 | 0.03 | 2.22E-07 | TRUE | MOG |
| rs11759438 | 6:169633335 | THBS2 | C/T | 0.5 | 0.1 | 0.02 | 2.94E-07 | TRUE | THBS2 |
| rs34763594 | 7:17878797 | SNX13 | G/A | 0.1 | -0.17 | 0.03 | 4.75E-08 | FALSE |  |

Results are shown for all variants with p-value < 1e-6 with a 3Mb window from a GWAS of ProtAgeGap in UKB Europeans (n=38,865). Variants that are cis-pQTLs for any of the 204 proteins in the original ProtAge clock ( $\pm 1$ Mb around the gene region for each protein) are indicated in the "ProtAge cis-pQTL" column, and if the variant is a cis-pQTL then the corresponding ProtAge proteins in-cis are indicated. EF: effect allele; OA: other allele; SE: standard error; pQTL: protein quantitative trait locus.

To investigate where in the genome the genetic contributions to ProtAgeGap originate, we performed partitioned heritability analysis using LDSC with the baseline LD v2.2 model (Table S1). This approach estimates the proportion of SNP heritability explained by various functional categories while adjusting for LD structure. We observed strong enrichment of heritability in evolutionarily conserved and functionally critical regions. Notably, SNPs annotated as protein-coding comprised only 1.4% of all SNPs (Prop._SNPs = 0.0143) but explained 47.5% of total SNP heritability (Prop._h2 = 0.475), corresponding to an enrichment of 33.3-fold (Enrichment = 33.25, p = 1.7 × 10⁻⁴). Similarly, conserved elements across vertebrates (Lindblad-Toh et al.) represented 2.6% of SNPs and accounted for 72.6% of heritability (Enrichment = 28.26, p = 2.4 × 10⁻⁵). Regions with high GERP++ RS scores, indicative of strong evolutionary constraint, were also markedly enriched. Specifically, SNPs in the top GERP score bin (RS > 4) accounted for 41.2-fold more heritability than expected (p = 5.4 × 10⁻⁴). Taken together, these results highlight that ProtAgeGap heritability is enriched in evolutionarily conserved, coding, and transcriptionally active genomic regions, underscoring the biological relevance of these features and pointing toward strong functional constraint and regulatory architecture in the genetic basis of proteomic aging.

### Gene set enrichment

Using the ProtAgeGap GWAS summary statistics, we mapped individual GWAS variants to genes and performed a gene-level analysis via MAGMA v1.10 using the 1000 genomes European LD reference data, aggregating individual variant effects to the level of whole genes and jointly testing each gene for its association with ProtAgeGap. We then performed a competitive gene set analysis using MSigDB gene sets from Kyoto Encyclopedia of Genes and Genomes (KEGG), REACTOME, and Gene Ontology (GO) biological processes (BP) and molecular function (MF) (Fig. 1c; Table S2). While no enrichments reached FDR < 0.05, we observed consistent biological themes returned as nominally significant across gene sets at marginal multiple testing correction levels (FDR ∼0.13–0.35). The pathways enriched at FDR < 0.2 converge on a few core themes: (1) early developmental programs (e.g., embryo development, in utero development, organ morphogenesis such as atrioventricular-node or bile-duct formation), (2) growth-factor signaling (e.g., IGF-binding proteins, Notch processing, growth hormone response, and GDNF/neurotrophin signaling), (3) epigenetic and post-translational regulation (lysine and histone methyltransferases), and (4) core metabolism and homeostasis (purine salvage, serine hydrolases, and nutrient-response regulators). These results underscore how fundamental developmental, signaling, and chromatin-modifying machinery intersect with metabolic control to shape proteomic aging.

### Genetic correlation with longevity and key aging-related traits

We calculated the genetic correlation between proteomic aging and key aging traits using LDSC (Fig. 1d). The results provide a clear genetic signature connecting proteomic aging to lifespan and developmental traits in both males and females. The strongest genetic correlation observed was with longevity^4^ (rg = −0.83; SE = 0.37; p-value = 0.025), confirming that proteomic aging is a trait that is strongly linked to longevity and maximum lifespan. Significant correlation was also observed with telomere length (rg = −0.16; SE = 0.07; p-value = 0.014), which is a biomarker of aging. While proteomic age was strongly genetically correlated with the intrinsic epigenetic age acceleration (IEAA)^5^ epigenetic clock (rg = 0.36; SE = 0.14; p-value = 0.011), no other epigenetic clocks (PhenoAge,^6^ GrimAge,^7^ Hannum^8^) showed significant genetic correlations. This confirms our previous work demonstrating that the underlying biology represented by our proteomic clock does not overlap strongly with existing epigenetic clocks.^2^

Genetic correlations of ProtAgeGap with BMI (rg = 0.14; SE = 0.05; p-value = 0.007), HDL cholesterol (rg = −0.11; SE = 0.05; p-value = 0.040), systolic blood pressure (rg = 0.16; SE = 0.05; p-value = 0.004), and C-reactive protein (CRP; rg = 0.14; SE = 0.05; p-value = 0.008) further demonstrate the genetic connections between proteomic aging and cardiometabolic health and inflammation.

Lastly, we observed strong genetic correlation between ProtAgeGap and several pubertal timing traits in females, including age at menarche (rg = −0.30; SE = 0.06; p-value = 1e-6), age at first sexual intercourse (rg =-0.18; SE = 0.07; p-value = 0.008), and age at first natural birth (rg = −0.18; SE = 0.07; p-value = 0.010). In males, there was a significant genetic correlation between ProtAgeGap and age at first facial hair (rg =-0.26; SE = 0.09; p-value = 0.006), but not age at first voice breaking. This underscores the genetic underpinnings linking proteomic aging to developmental and pubertal timing earlier in life.

### FTO locus as a potential key mechanism in aging-related antagonistic pleiotropy

We further used Bayes factor colocalization via the coloc^9^ package in R to assess whether fine-mapped loci from the ProtAgeGap GWAS share causal variants with fine-mapped loci for traits related to childhood growth and development, sex and growth hormones, and adulthood cardiometabolic disease. We observed significant colocalization (H4 posterior probabilities > 0.80) at the same intronic variant in the *FTO* locus (rs11642015) between ProtAgeGap, age at menarche, and adulthood measures of sex hormone-binding globulin (SHBG), Insulin-like Growth Factor 1 (IGF-1), HDL cholesterol, obesity, and type 2 diabetes (T2D) (Fig. 2). These results demonstrate a common genetic underpinning in the *FTO* locus between these early life development and adulthood cardiometabolic traits, and implicate proteomic aging as a key axis that unites these traits in a phenotype of accelerated biological aging that spans the life course.

**Fig. 2.**
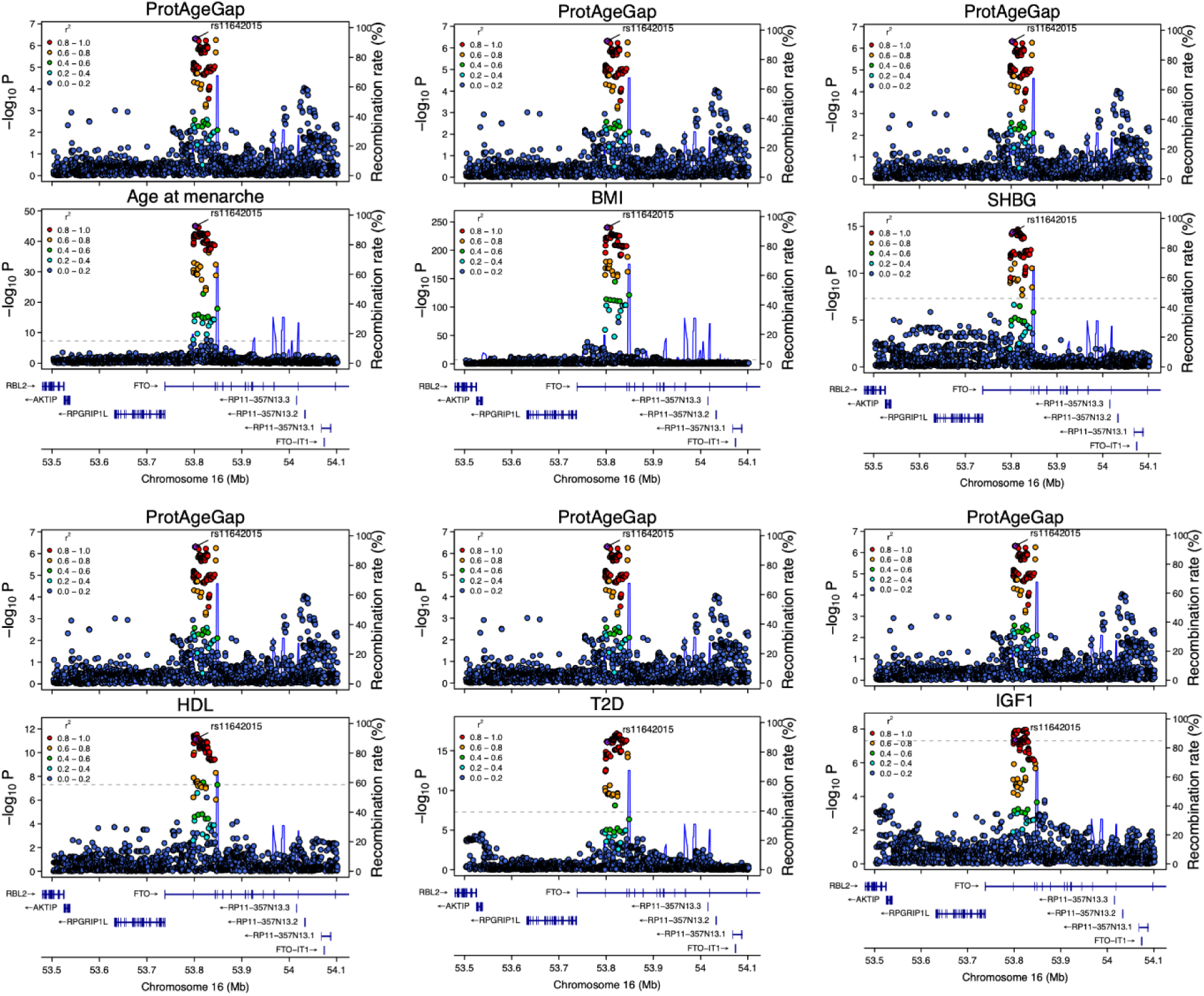
Genetic colocalization between proteomic aging and aging-related traits. Genetic colocalization of a fine-mapped region in the *FTO* gene. Fine-mapping for each trait was performed using GWAS summary statistics and LD matrices calculated from UK Biobank Europeans (n=38,865 for ProtAgeGap; n=361,194 for all other traits). Fine-mapping in each trait was performed using SUSiE and then colocalization performed using approximate Bayes factor colocalization. BMI: body mass index; SHBG: sex hormone binding globulin; HDL; high density lipoprotein; T2D: type 2 diabetes: IGF1: insulin-like growth factor 1; LD: linkage disequilibrium.

### ProtAgeGap polygenic scores predict numerous traits across the life course

Using the ProtAgeGap GWAS summary statistics from the UKB, we developed PGS for ProtAgeGap using genotyping data available in FinnGen (n=500,348), China Kadoorie Biobank (CKB; n=100,640), European participants from All of Us (n=117,415), and the ABCD Study (n=5,204). In the subset of FinnGen and CKB participants who had both genetic and proteomic data available (n=1,990) the ProtAgeGap PGS showed modest performance in predicting the actual ProtAgeGap proteomic phenotype (R^2^ = 0.006 and R^2^ = 0.008, respectively). The ABCD study and All of Us did not have proteomic data available for direct calculation of ProtAgeGap.

Within FinnGen, we found that the ProtAgeGap PGS meaningfully predicts lifespan and age at death. Specifically, among the FinnGen participants who died during follow up (n=60,914), those in the bottom quintile of the ProtAgeGap PGS were 43% more likely to have an age at death > 95 years, compared to those within the top quintile of the PGS (OR: 1.43; 95% CI: 1.12-1.84; p-value: 0.005).

We further carried out a phenome-wide association study (PheWAS) in FinnGen and the All of Us Study, testing the association of ProtAgeGap PGS with 3,521 and 1,702 clinical and medication endpoints in FinnGen and All of Us, respectively (Fig. 3). Among these, 264 traits showed a significant association with ProtAgeGap PGS below the FDR threshold of 0.05 in FinnGen (Fig. 3a), and 40 traits had FDR-significant associations in All of Us (Fig. 3b). ProtAgeGap PGS associations that replicated across both PheWAS include obesity, hypertension, hypercholesterolemia, hyperlipidemia, heart disease, type 2 diabetes, emphysema, spondylosis without myelopathy (wear & tear on spine), and osteoarthritis (Table 4). Further, in the CKB we carried out additional association testing of the ProtAgeGap PGS in relation to select clinical risk factors and disease outcomes. We further replicated associations between ProtAgeGap PGS and obesity, hypertension, heart disease, and osteoarthritis in this cohort, despite testing them in participants from a different genetic ancestry (east Asian) compared to our GWAS population (European).

**Fig. 3.**
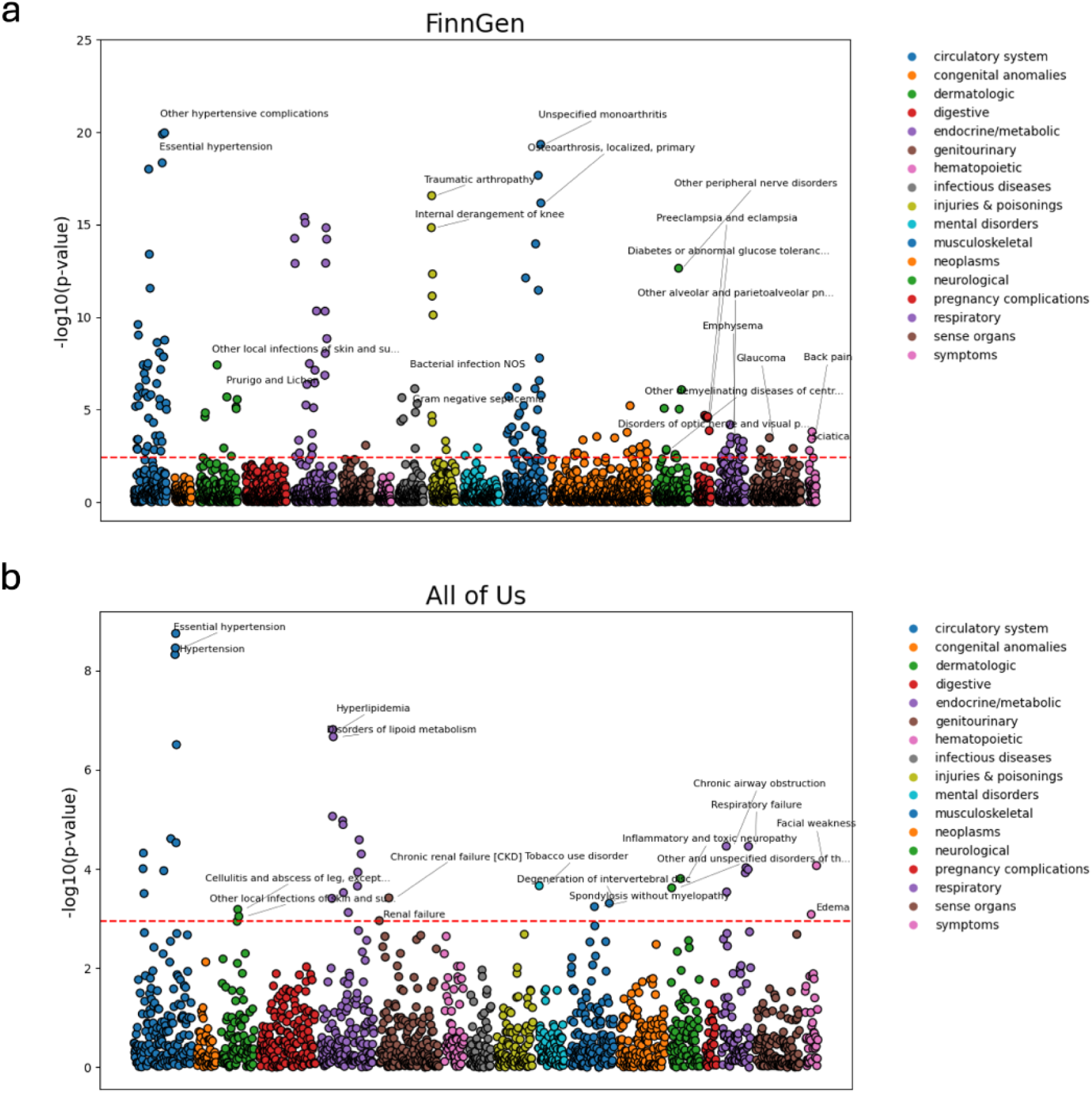
Proteomic aging PGS PheWAS associations in FinnGen and All of Us. **a)** PheWAS of associations between the ProtAgeGap PGS and 3,521 clinical endpoints in FinnGen (n=500,348). Models were logistic regressions adjusted for age, sex, genotyping chip, cohort, and 10 genetic principal components. b) PheWAS of associations between the ProtAgeGap PGS and 1,702 clinical endpoints in All of Us (n=117,415). Models were logistic regressions adjusted for age, sex, and 10 genetic principal components. In both **a)** and **b)**, the red dotted line corresponds to the 5% FDR significance threshold, however only the top two hits per each category of clinical endpoints is labelled for the sake of space. PheWAS: phenome-wide association study; PGS: polygenic score; FDR: false discovery rate.

**Table 4.** Replicated ProtAgeGap PGS and disease associations across biobanks.

|  |  |  |  |  |  |  |  |  |  |  |
| --- | --- | --- | --- | --- | --- | --- | --- | --- | --- | --- |
| Phecode | Endpoint | Category | FinnGen (n=500,348) |  |  |  | All of Us (n=117,415) |  |  |  |
|  |  |  | Beta | CI_low | CI_high | P-value | Beta | CI_low | CI_high | P-value |
| 250.2 | Type 2 diabetes | endocrine/<br>metabolic | 0.03 | 0.02 | 0.04 | 4.11E-16 | 0.04 | 0.02 | 0.06 | 2.57E-05 |
| 272.1 | Hyperlipidemia | endocrine/<br>metabolic | 0.02 | 0.02 | 0.03 | 7.32E-08 | 0.04 | 0.03 | 0.06 | 1.55E-07 |
| 272.11 | Hypercholesterolemia | endocrine/<br>metabolic | 0.03 | 0.02 | 0.04 | 3.59E-07 | 0.04 | 0.02 | 0.06 | 8.67E-06 |
| 278 | **Overweight, obesity**<br>**and other**<br>**hyperalimentation** | endocrine/<br>metabolic | 0.04 | 0.03 | 0.05 | 4.69E-11 | 0.03 | 0.01 | 0.04 | 2.99E-04 |
| 278.1 | **Obesity** | endocrine/<br>metabolic | 0.04 | 0.03 | 0.05 | 4.79E-11 | 0.03 | 0.02 | 0.05 | 1.28E-05 |
| 401.1 | **Essential**<br>**hypertension** | circulatory<br>system | 0.03 | 0.03 | 0.04 | 4.62E-19 | 0.04 | 0.03 | 0.06 | 3.52E-09 |
| 401.2 | **Hypertensive heart**<br>**and/or renal disease** | circulatory<br>system | 0.05 | 0.04 | 0.07 | 1.35E-08 | 0.08 | 0.05 | 0.11 | 1.80E-09 |
| 401.21 | **Hypertensive heart**<br>**disease** | circulatory<br>system | 0.06 | 0.04 | 0.08 | 1.40E-08 | 0.07 | 0.04 | 0.1 | 2.95E-05 |
| 496.1 | Emphysema | respiratory | 0.03 | 0.01 | 0.04 | 3.02E-04 | 0.07 | 0.03 | 0.11 | 2.90E-04 |
| 502 | Postinflammatory<br>pulmonary fibrosis | respiratory | 0.07 | 0.03 | 0.11 | 1.03E-03 | 0.09 | 0.05 | 0.14 | 1.19E-04 |
| 504 | Other alveolar and<br>parietoalveolar<br>pneumonopathy | respiratory | 0.06 | 0.03 | 0.09 | 6.42E-05 | 0.11 | 0.06 | 0.17 | 9.39E-05 |
| 686 | Other local infections<br>of skin and<br>subcutaneous tissue | dermatolog<br>ic | 0.03 | 0.02 | 0.05 | 3.82E-08 | 0.05 | 0.02 | 0.08 | 9.07E-04 |
| 721.1 | **Spondylosis without**<br>**myelopathy\*** | musculosk<br>eletal | 0.02 | 0.01 | 0.04 | 4.43E-04 | 0.03 | 0.01 | 0.05 | 5.73E-04 |

Shown are phecodes significantly associated with the ProtAgeGap PGS after FDR correction in both the FinnGen and All of Us cohorts. Endpoints shown in bold are those that were also associated with the ProtAgeGap PGS in the CKB (n=100,640). \*Spondylosis was not available in the CKB, but the ProtAgeGap was associated with a closely related trait - osteoarthritis. ProtAgeGap: proteomic age gap; PGS: polygenic score; FDR: false discovery rate.

Within the ABCD Study, we tested the associations of the ProtAgeGap PGS with weight, height, puberty timing, brain volume, and cognition among children aged 9-18 years old. In mixed effects models accounting for repeat measurement of developmental traits across all study visits, we observed significant associations between the ProtAgeGap PGS and a derived developmental stage score, weight, height, BMI, and pubertal stage, but we observed no significant associations with brain volume or cognition (Fig. 4a). This indicates that ProtAgeGap likely represents an underlying life course phenotype that begins in childhood and that this trajectory can be detected using genomic data in children as early as 9 years old. Performing the same regressions among each study wave separately (baseline, 9-11 years; year 2, 11-13 years; year 4, 13-15 years; year 6, 15-18 years) revealed stronger associations in earlier windows of life, with many associations not significant once participants reached 11-15 years of age (Fig. 4b). These results point to a more critical period before 11 years of age in childhood and adolescence when genetic risk for proteomic aging seems to have a much stronger association with puberty and developmental traits. Summary statistics from all PGS PheWAS and association testing can be found in Tables S3-S4.

**Fig. 4.**
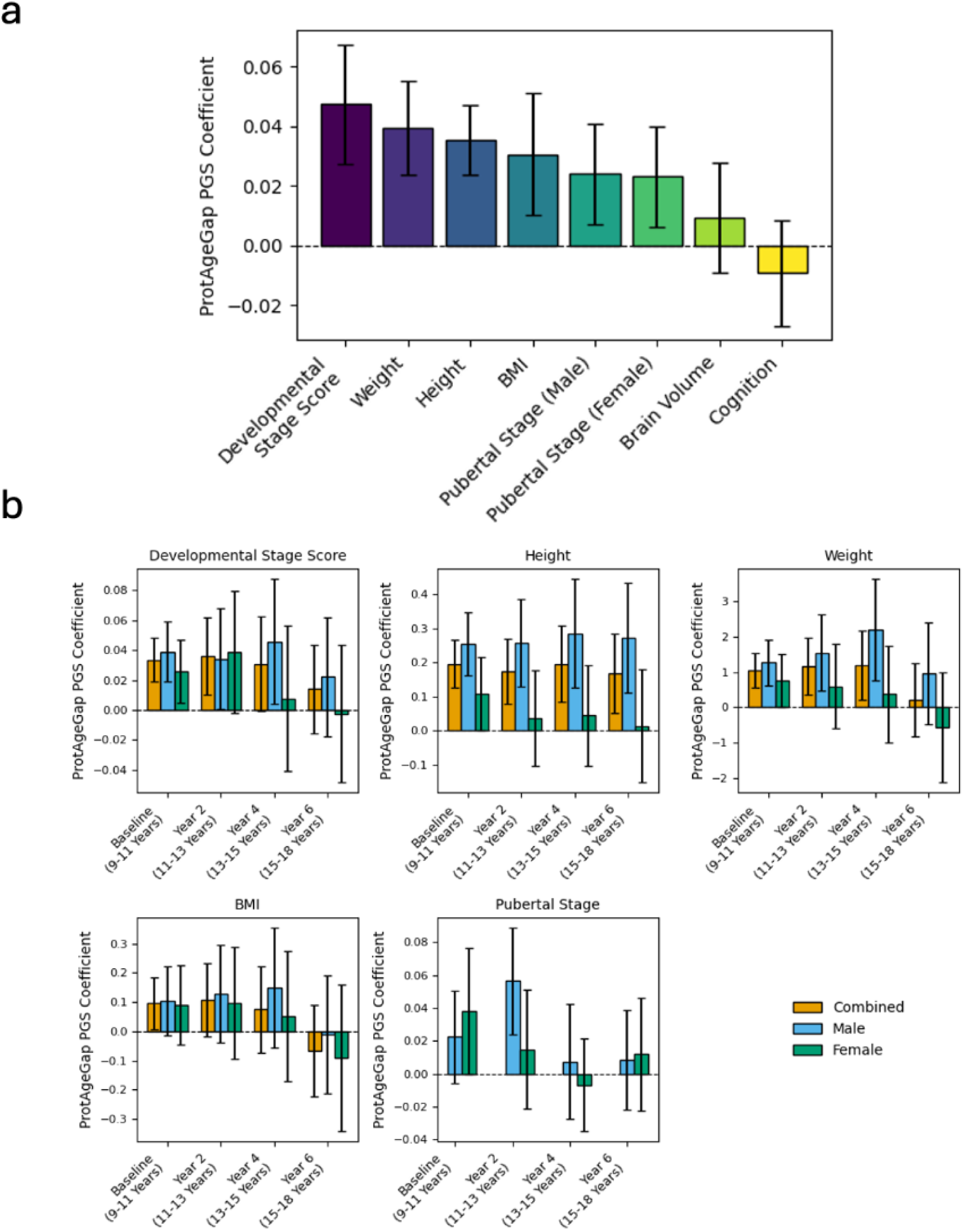
Proteomic aging PGS associations with growth and puberty in the ABCD Study. **a)** Mixed effect linear regression models between the ProtAgeGap PGS and developmental traits in childhood participants from the ABCD study (n=5,204) aged 9-18 years old. All regressions were adjusted for age, age^2^, sex, study site, and 10 genetic principal components, with a random intercept fitted for participant identifier to account for repeated outcome measurements across the three study visits for each participant. **b)** Linear regression models assessing the effect of the ProtAgeGap PGS on each developmental outcome in ABCD study participants, with a separate regression fitted within each study visit separately. PGS: polygenic score.

## Discussion

In this study, we present one of the first genome-wide association analyses of proteomic aging, leveraging plasma proteomic profiles and genomic data from nearly 39,000 individuals of European ancestry in the UKB. Our findings illuminate the genetic underpinnings of ProtAgeGap—a complex proteomic phenotype capturing deviations between biological and chronological age—and reveal shared genetic architecture of ProtAgeGap with longevity, growth and developmental biology, and cardiometabolic and musculoskeletal diseases.

Our work provides a new framework for understanding the biology of proteomic aging as a lifelong process. Proteomic aging captures dynamic physiological changes shaped by regulatory and evolutionarily constrained regions of the genome. Its polygenic architecture overlaps with early-life growth and development, adulthood morbidity and chronic disease, and longevity, highlighting a continuous biological axis that spans across the life course. Our results also reinforce the value of multi-omics aging clocks as tools for discovering conserved, genetically encoded aging pathways.

While biological age clocks have been frequently associated with all-cause and premature mortality across a number of studies,^10,11^ the evidence linking biological aging clocks directly to longevity has so far been much more sparse (with a few notable exceptions^12,13^). Our work provides evidence of a direct genetic link between proteomic aging and longevity. We observed an extremely strong genetic correlation (rg: −0.83) between proteomic aging and longevity, and in FinnGen those in the top quintile of our ProtAgeGap PGS had significantly higher odds of living to at least 95 years of age. Furthermore, the ProtAgeGap PGS showed broad associations with over 250 phenotypes, including obesity; cardiometabolic, lung, and musculoskeletal diseases; and related medication usage—highlighting its relevance to adult chronic disease burden. These results further reinforce our previous demonstration that proteomic aging is likely involved in age-related multimorbidity and mortality,^2^ and confirm that the molecular phenotype captured by our proteomic age clock is linked to not just healthspan but also lifespan and longevity.

Puberty and developmental timing traits such as age at menarche and childhood BMI have previously been associated with adulthood obesity and cardiometabolic disease risk.^14,15^ However, our work provides new evidence of a genetic link between adulthood proteomic aging and these early life developmental traits through multiple lines of evidence, including: (1) strong demonstrated genetic correlation of proteomic aging with age at menarche, and colocalization between these two traits at a causal variant in the FTO locus; (2) strong enrichment for early life developmental processes in our gene set enrichment analysis; and (3) observed associations between the ProtAgeGap PGS and childhood weight, height, developmental stage, and puberty timing in participants aged 9-18 years old from the ABCD Study. Interestingly, we observed no genetic correlation with total number of births, and we also observed far fewer genetic variants that were associated with both adulthood height and ProtAgeGap compared with childhood height and ProtAgeGap. These results indicate that ProtAgeGap is less involved in overall reproductive activity and final height achieved, and is rather related to the timing and trajectory of growth and puberty early in life.

Taken together, these results provide evidence that the biological underpinnings of aging have diverse influences across the life course, influencing early life growth and development, adulthood disease risk, and overall propensity for longevity. In particular, our results suggest that accelerated biological aging is a lifelong phenotype, whereby the same genetic underpinnings contribute to earlier growth and development, higher risk of adulthood obesity and chronic disease, and higher risk of dying younger. These diverse connections may reflect a central role of growth and obesity in aging across the life course, which is reflected in our results demonstrating strong associations of the ProtAgeGap PGS with obesity and cardiometabolic traits, as well as genetic colocalization observed in the FTO locus across traits. In mice, it has been shown that FTO overexpression leads to earlier puberty onset,^16^ and numerous Mendelian randomization studies have shown a likely causal role of early life obesity on earlier age at menarche.^17–19^ Previously published and robust evidence on the role of obesity on lifespan^20,21^ and disease multimorbidity^22,23^ further support this interpretation of the central role of obesity in aging across the life course.

Interestingly, our results reinforce the conclusions that current proteomic age clocks and epigenetic age clocks represent different and largely non-overlapping biological pathways and mechanisms. In our previous work, we demonstrated that our proteomic aging clock includes few proteins corresponding to genes from epigenetic age clocks.^2^ Our current analyses also demonstrate poor genetic correlation between ProtAgeGap and most epigenetic clocks. This may explain why epigenetic clocks are associated with different phenotypes and are less robustly associated with development and growth. For example, previous work has shown that epigenetic age deviation has been associated with age at menopause but not menarche.^24^

Several limitations warrant consideration. First, the discovery cohort was restricted to individuals of European ancestry, limiting generalizability across diverse populations. While we replicated many associations in cohorts from Asia and the U.S., future efforts should expand these analyses to more ancestrally diverse populations. Second, while we detected significant genetic correlations and PGS-trait associations, functional validation is needed to confirm causality and elucidate biological mechanisms. Integration with transcriptomic and proteomic QTLs may help prioritize causal genes, and experimental studies could further validate candidate pathways. Lastly, although the PGS was predictive across cohorts, the variance in ProtAgeGap explained by the PGS was modest, reflecting the context-dependent and environmentally driven nature of aging.^25^

In conclusion, this study reveals that proteomic aging is genetically influenced, evolutionarily constrained, and interconnected with human development, disease, and longevity. By mapping the genomic architecture of proteomic age, we uncover shared biological pathways that link early-life growth to late-life health outcomes—offering a window into the lifelong molecular mechanisms that shape human aging.

## Methods

### Cohorts

The UKB is a prospective cohort study with extensive genetic and phenotype data available for 502,505 individuals residing in the United Kingdom who were recruited between 2006 and 2010. The full UKB protocol is available online (https://www.ukbiobank.ac.uk/media/gnkeyh2q/study-rationale.pdf). We restricted our UKB sample to those participants with Olink Explore data available at baseline who were randomly sampled from the main UKB population (n = 45,445).

The CKB is a prospective cohort study of 512,724 adults aged 30–79 years who were recruited from ten geographically diverse (five rural and five urban) areas across China between 2004 and 2008. Details on the CKB study design and methods have been previously reported.^26^ We restricted our CKB sample to those participants with genotyping and clinical data, and who were genetically unrelated to each other (n=100,640).

The FinnGen study is a public–private partnership research project that has collected and analyzed genome and health data from 500,000 Finnish biobank donors to understand the genetic basis of diseases.^27^ FinnGen includes nine Finnish biobanks, research institutes, universities and university hospitals, 13 international pharmaceutical industry partners and the Finnish Biobank Cooperative (FINBB). The project utilizes data from the nationwide longitudinal health register collected since 1969 from every resident in Finland. In FinnGen, we restricted our analyses to those participants with genotyping and clinical data (n=500,348).

The All of Us cohort aims to engage a longitudinal cohort of one million or more US participants, with a focus on including populations that have historically been under-represented in biomedical research. Details of the All of Us cohort have been described previously.^28^ Briefly, the primary objective is to build a robust research resource that can facilitate the exploration of biological, clinical, social and environmental determinants of health and disease. Health data are obtained through the electronic medical record and through participant surveys. Survey templates can be found on our public website: https://www.researchallofus.org/data-tools/survey-explorer/. We restricted our analyses to participants who have genotyping and clinical record data available (n=117,415).

The ABCD study is a longitudinal study across 21 data acquisition sites in the United States following 11,880 children starting at 9–11 years. This study uses 6.0 data from the NIH Brain Development Cohorts archive (nbdc-datahub.org) (DOI: https://doi.org/10.82525/jy7n-g441). The ABCD cohort was recruited to ensure the sample was as close to a national representative as possible, and therefore exhibits large sociodemographic diversity.^29^ There is an embedded twin cohort and many siblings. As the discovery cohort for GWAS results were performed in individuals of European ancestry, the analyses were conducted in participants of European ancestry, with sample sizes as follows: baseline (ages 9–11), 6,359; 2-year follow-up (ages 11–13), 4,706; 4-year follow-up (ages 13–15), 3,560; and 6-year follow-up (ages 15–18), 2,544.

### ProtAgeGap score

ProtAgeGap was developed and calculated previously in our UKB dataset, with extensive details on methods reported in our previous publication.^2^ Briefly, to calculate proteomic age we first split all UKB participants with available plasma proteomics data (n = 45,441) into 70:30 train–test splits. In the training data (n = 31,808), we trained a model to predict age at recruitment using all 2,897 proteins in a single LightGBM^30^ model using fivefold cross-validation. We then carried out Boruta^31^ feature selection to identify a subset of 204 proteins that predict age above random noise. Finally, we re-tuned model hyperparameters for a new model with the subset of selected proteins using the same procedure as before. Both tuned LightGBM models before and after feature selection were checked for overfitting and validated by performing fivefold cross-validation in the combined train set and testing the performance of the model against the holdout UKB test set. Once the final model with Boruta-selected proteins was trained in the UKB, we calculated protein-predicted age (ProtAge) for the entire UKB cohort (n = 45,441) using fivefold cross-validation. Within each fold, a LightGBM model was trained using the final hyperparameters and predicted age values were generated for the test set of that fold. We then combined the predicted age values from each of the folds to create a measure of ProtAge for the entire sample. ProtAgeGap was then calculated by taking the difference in years between ProtAge and age a recruitment for each participant.

### Genetic QC and GWAS

Genetic ancestry was mapped using previously calculated genetic ancestry in our UKB data.^3^ For genetics analyses, we removed all participants not of European ancestry and performed sample quality control (QC) by removing UKB individuals with discordant self-reported and genetic sex (field IDs 31, 22001), with sex chromosome aneuploidy (field ID 22019), with ten or more third-degree relatives identified (field ID 22021), and those marked as outliers for heterozygosity and missing rates (field ID 22027). Variant QC was performed by removing variants with call rate lower than 95%, MAF < 0.01, and Hardy Weinberg equilibrium < 1e-6.

We carried out a genetic principal components analysis (PCA) within the final study sample using Hail. Before running the PCA, we first removed the MHC region (6:28477797 - 6:33448354; hg37) and regions of long-range high linkage disequilibrium (LD) (https://genome.sph.umich.edu/wiki/Regions_of_high_linkage_disequilibrium_(LD)) and then LD pruned using an r^2^ threshold of 0.2 in 5kb windows.

GWAS was carried out using the linear_regression_rows() function in Hail on the UKB v3 imputed genotype dosages (hg37) across all autosomes, adjusted for age, age^2^, sex, age*sex, age^2^*sex, genotype array, and 20 genetic principal components calculated in our sample. Gene names were mapped to variants in the GWAS summary statistics using the Ensembl Variant Effect Predictor (VEP) in Hail.

### Downstream genetic analyses

Genetic correlation and SNP heritability estimation was carried out using GWAS summary statistics with the MHC region removed using the ldsc python package and using LD reference information from the 1000 genomes project European samples. GWAS summary statistics used for genetic correlation analyses included: longevity^4^, epigenetic clocks,^32^, number of children ever born,^33^ age at first sexual intercourse,^34^ age at first natural birth,^34^ telomere length.^35^ Summary statistics for age at first facial hair, age at voice breaking, and age a menopause were taken from Karczewski et al. (2024).^36^ Summary statistics for all other traits used in genetic correlation analyses were taken from Kanai et al. (2021).^37^ For fine-mapping and colocalization analyses, an in-sample LD matrix was computed in Hail with a 10Mb radius and all covariates included in the GWAS. Fine-mapping was conducted using Sum of Single Effects” (SuSiE)^38^ using the susieR package in R. Within each 3Mb region around a lead variant with a p-value < 1e-6, we fine-mapped variants using a within-sample LD matrix and credible set coverage set to 95%. We performed colocalization analyses within each region for which a credible set was found in our initial ProtAgeGap GWAS using approximate Bayes factors using the coloc.bf_bf function in the coloc R package. GWAS summary statistics and SuSiE fine-mapping results for other non-ProtAgeGap traits used for colocalization analyses were taken from a previous GWAS and fine-mapping project conducted in UKB Europeans (n=361,194).^37^

### Developmental staging score in the ABCD Study

In an approach similar to the ProtAge model we trained a model to predict chronological age from a range of anthropomorphic, neurocognitive, pubertal staging, and magnetic resonance brain imaging (details found here^39,40^) using a light gradient-boosting machine model using LightGBM (v 4.5.0, in python v3.11.10). The total number of features in this model totaled 13,848 brain imaging and 82 non-brain imaging variables. To reduce the dimensionality of this dataset we performed principle component analysis to reduce the brain imaging variables to the top 10 components ultimately resulting in 92 features input for LightGBM. We fitted models separately in: a) individuals assigned female and male at birth, and b) stratified by wave of the study (e.g. baseline (9-11 year old), year 2 (11-13 years old), year 4 (13-15 year olds), and year 6 (15-18 years old)). Models were fit cross-validated (5-fold) ensuring that genetically related individuals were not split across training and testing folds. For the LightGBM model we use the following parameters: num_leaves=50, learning rate=0.1, min_data_leaf=20, lambda_l1=0, and lambda=0. Across all time points both female and male models achieved high performance at R^2^=0.85. The difference between this model’s predicted age and chronological age is what we refer to as the “Developmental Staging Score”, which indicates accelerated or decelerated development with reference to chronological age.

### Polygenic score analyses

In all cohorts, PGS were calculated using PRS-cs^41^ based on the UKB ProtAgeGap GWAS summary statistics. The MHC region was removed from summary statistics prior to calculating the ProtAgeGap PGS in each cohort. In FinnGen and All of Us, we carried out a phenome-wide association study (PheWAS) in the full set of participants with genotyping data (FinnGen n=500,348; All of Us n=117,415) each cohort by regressing ProtAgeGap PGS against all available endpoints using logistic regression adjusted for age, sex, 10 genetic PCs, and additionally for genotyping chip and cohort in FinnGen only. We calculated the false discovery rate (FDR) separately for each cohort using the benjamini-hochberg method.^42^ In FinnGen, we used all centrally available R12 endpoints derived from ICD-9 and ICD-10 codes using linked electronic health records (EHR), keeping endpoints with at least 100 cases (3,521 endpoints). In All of Us, we used centrally coded endpoints using phecodes derived from ICD-9 and ICD-10 codes extracted from EHRs, again limiting to endpoints with at least 100 cases. In both FinnGen and All of Us, phecodes were systematically mapped to descriptive phecodes as documented in the PheWAS Catalog (https://phewascatalog.org/). For each phecode, individuals were categorized as a case if they had at least one corresponding ICD code associated with it.

We estimated the phenotypic variance in proteomic aging explained by the ProtAgeGap PGS in FinnGen and CKB using linear regression. We first regressed ProtAgeGap onto basic covariates (CKB: age, sex, region code, and 11 genetic PCs; FinnGen: age, sex, genotyping chip, cohort, 10 genetic PCs) and retrieved the R^2^ estimate. We then fitted a second model with the same covariates plus the ProtAgeGap PGS, retrieved the R^2^ estimate from this second model and then subtracted the R^2^ estimate from model 1 to calculate the phenotypic variance explained by the ProtAgeGap PGS only.

For PheWAS, we serially regressed each endpoint from FinnGen and All of Us onto the ProtAgeGap PGS via binomial logistic regression using slightly differing sets of covariates in each cohort (FinnGen: age, sex, genotyping chip, cohort, and 10 genetic PCs; All of Us: age, sex, and 10 genetic PCs). Within each PheWAS separately, we performed FDR adjustment using the Benjamini-Hochberg method.

For the ABCD cohort analyses we used TOPMED imputed genetic data from baseline blood or saliva samples to generate PGS, details of genetic data are described elsewhere.^43^ For phenotypic associations with PGS, we used mixed effect linear regression models between the ProtAgeGap PGS and developmental traits in childhood participants from the ABCD study (n=5,204) aged 9-18 years old. These developmental traits included: a “developmental stage score” (details described above), weight, height, body mass index, total brain volume (), cognition (NIH cognition toolbox fluid composite summary measure: http://www.nihtoolbox.org ^44^), and a 5 point categorical pubertal staging assessment for each sex.^44,45^ All regressions adjusted for age, sex assigned at birth, study site, and 10 genetic principal components, with a random intercept fitted for participant identifier nested within genetic family membership to account for repeated outcome measurements across the four study visits for each participant as well as the high degree of relatedness within the study. For the regression model predicting total brain volume we additionally included a fixed effect for the magnetic resonance imaging scanner id. In the ABCD Study, we also calculated linear regression models assessing the effect of the baseline ProtAgeGap PGS on each developmental outcome in ABCD study participants, with a separate regression fitted within each of the study visits.

## Acknowledgments

We gratefully acknowledge all participants and research teams involved in UKB, FinnGen, CKB, and the ABCD Study. Data used in the preparation of this article were obtained from the Adolescent Brain Cognitive Development (ABCD) Study (https://abcdstudy.org), held in the NIH Brain Development Cohorts (NBDC) data hub (nbdc-datahub.org). This is a multisite, longitudinal study designed to recruit more than 10,000 children aged 9-10 and follow them over 10 years into early adulthood. The ABCD Study is supported by the National Institutes of Health and additional federal partners under award numbers U01DA041022, U01DA041028, U01DA041048, U01DA041089, U01DA041106, U01DA041117, U01DA041120, U01DA041134, U01DA041148, U01DA041156, U01DA041174, U24DA041123, U24DA041147, U01DA041093, and U01DA041025. A full list of supporters is available at https://abcdstudy.org/federal-partners.html. ABCD consortium investigators designed and implemented the study and/or provided data but did not all necessarily participate in analysis or writing of this report. This manuscript reflects the views of the authors and may not reflect the opinions or views of the NIH or ABCD consortium investigators. The ABCD data repository grows and changes over time. The data were downloaded from nbdc-datahub.org as the 6.0 data release (DOI: https://doi.org/10.82525/jy7n-g441).

## Data availability

GWAS summary statistics will be made available in a public repository upon publication of this manuscript. UKB data are available through a procedure described at https://www.ukbiobank.ac.uk/enable-your-research. The CKB is a global resource for the investigation of lifestyle, environmental, blood biochemical and genetic factors as determinants of common diseases. The CKB study group is committed to making the cohort data available to the scientific community in China, the United Kingdom and worldwide to advance knowledge about the causes, prevention and treatment of disease. For detailed information on what data are currently available to open-access users and how to apply for them, please visit https://www.ckbiobank.org/data-access. A research proposal will be requested to ensure that any analysis is performed by bona fide researchers. Researchers who are interested in obtaining additional information or data that underline this paper should contact. For any data that are not currently available via open access, researchers may need to develop a formal collaboration with the CKB study group.

FinnGen as a research project is granted use of national healthcare data and biospecimens according to national and European regulations (GDPR), which preclude the research project from distributing individual level data. However, any researcher can apply for the health register data from the Finnish Data Authority Findata (https://findata.fi/en/permits/) and for all FinnGen individual-level genotype data (and other profiling data) generated by the project from Finnish biobanks via the Fingenious portal (https://site.fingenious.fi/en/) hosted by the Finnish Biobank Cooperative FINBB (https://finbb.fi/en/). Summary statistics from all FinnGen analyses are available publicly - more details about accessing other FinnGen results can be found here: https://www.finngen.fi/en/access_results. Academic users wishing to work with the FinnGen project resource directly can contact us through a [web form](https://docs.google.com/forms/d/1IeeLPPy0ekFgZQomjYvqdHZa420UuA4zQ7eMgMASCYs/edit). Learn more about collaboration opportunities at: https://www.finngen.fi/en/how-we-collaborate.

Access to individual-level data from the All of Us research program is available to researchers whose institution has signed a data use agreement with All of Us (https://www.researchallofus.org/register/). All of Us provides a publicly available data browser (https://databrowser.researchallofus.org/) containing aggregate-level participant data for users to explore the available data, including genomic variants. Electronic health records (EHR) data, used for phenotyping, belongs to the registered tier dataset. Whole-genome sequencing data belongs to the controlled tier dataset, which requires additional training to access.

## Code availability

All code to conduct analyses and generate figures will be made available on GitHub at the time of publication of the final manuscript.

## Ethics approval

UKB data use (project application no. 31063) was approved by the UKB according to their established access procedures. UKB has approval from the North West Multi-centre Research Ethics Committee as a research tissue bank and as such researchers using UKB data do not require separate ethical clearance and can operate under the research tissue bank approval. The CKB complies with all the required ethical standards for medical research on human participants. Ethical approvals were granted and have been maintained by the relevant institutional ethical research committees in the United Kingdom and China. Study participants in FinnGen provided informed consent for biobank research, based on the Finnish Biobank Act. The FinnGen study is approved by the Finnish Institute for Health and Welfare (permit nos. THL/2031/6.02.00/2017, THL/1101/5.05.00/2017, THL/341/6.02.00/2018, THL/2222/6.02.00/2018, THL/283/6.02.00/2019, THL/1721/5.05.00/2019 and THL/1524/5.05.00/2020), Digital and Population Data Service Agency (permit nos. VRK43431/2017-3, VRK/6909/2018-3 and VRK/4415/2019-3), the Social Insurance Institution (permit nos. KELA 58/522/2017, KELA 131/522/2018, KELA 70/522/2019, KELA 98/522/2019, KELA 134/522/2019, KELA 138/522/2019, KELA 2/522/2020 and KELA 16/522/2020), Findata (permit nos. THL/2364/14.02/2020, THL/4055/14.06.00/2020, THL/3433/14.06.00/2020, THL/4432/14.06/2020, THL/5189/14.06/2020, THL/5894/14.06.00/2020, THL/6619/14.06.00/2020, THL/209/14.06.00/2021, THL/688/14.06.00/2021, THL/1284/14.06.00/2021, THL/1965/14.06.00/2021, THL/5546/14.02.00/2020, THL/2658/14.06.00/2021 and THL/4235/14.06.00/2021), Statistics Finland (permit nos. TK-53-1041-17 and TK/143/07.03.00/2020 (previously TK-53-90-20) TK/1735/07.03.00/2021 and TK/3112/07.03.00/2021) and Finnish Registry for Kidney Diseases permission/extract from the meeting minutes on 4 July 2019.

